# Efficacy of high-pressure balloon for the treatment of arteriovenous fistula stenosis: a meta-analysis

**DOI:** 10.1101/2021.03.08.21253176

**Authors:** Yu Li, Wenhao Cui, Jukun Wang, Chao Zhang, Tao Luo

## Abstract

**Objective:** The objective of the present study was to compare the effectiveness of high-pressure balloon (HPB) versus conventional balloon angioplasty (BA) in treating arteriovenous fistula (AVF) stenosis.

**Materials and Methods:** A meta-analysis was conducted using data acquired from PubMed, EMBASE, the Cochrane Library, SinoMed, CNKI, WanFang and VIP databases from the time the databases were established to November 2020. All analyses included in the studies comprised the subgroups of HPB and BA. The patency of AVF was compared between the two groups at 3 months, 6 months and 12 months after operation.

**Results:** Nine studies comprising 475 patients were included in the meta-analysis. The pooled results revealed that stenosis rate of AVFs treated with HPB was significantly lower than that of AVFs treated with conventional balloon at 3 months (OR= 0.37, 95% CI 0.21 to 0.67, p<0.001) and 6 months after operation (OR= 0.33, 95% CI 0.15 to 0.75, p=0.008). In addition, the technical success rate of HPB groups was high (OR= 0.14, 95% CI 0.05 to 0.35, p<0.001). However, no significant difference was observed between the experimental and control groups at 12 months after operation (OR= 0.61, 95% CI 0.29 to 1.25, p=0.18). No significant publication bias was observed in the analyses.

**Conclusion:** HPB is a potential primary option for the treatment of AVF stenosis, with a lower 3- and 6-month stenosis rate than BA. However, the long-term effect of HPB was not satisfactory; therefore, further research should be conducted to elucidate the relationship between the two groups.

## Introduction

Vascular access is the lifeline of patients with maintenance hemodialysis. Timely establishment and maintenance of the function of vascular access is crucial for the survival of such patients [1]. Arteriovenous fistula (AVF) has been in use for more than half a century since its first introduction in 1966. Considerable progress has been made in materials, surgical techniques and nursing measures associated with AVF, which has become the first choice of vascular access. However, stenosis of AVF and the resulting failure limit its use. Currently, AVF stenosis can be treated by surgery or through endovascular therapy. Endovascular therapy has become a key treatment because it confers numerous advantages such as minimal invasiveness and influence on dialysis rhythms. There are several types of balloons, including drug coated balloon (DCB), drug-eluting balloon (DEB) and high-pressure balloon (HPB). Different studies have presented varying evaluation results on the efficacy of different balloons for the treatment of AVF stenosis. Previous studies have demonstrated that DCB is superior to conventional balloon (CB) in terms of stenosis of AVFs, arteriovenous grafts (AVGs), and central venous catheters (CVC) [2–4]. However, studies comparing HPB and CB efficacies are scarce. Therefore, an updated meta-analysis was conducted to investigate the clinical effectiveness of HPB against CB for patients with AVF stenosis.

## Materials and Methods

### Selection criteria

① Studies included in the present meta-analysis focused on the treatment of AVF stenosis up to November 2020.
② Research methods were retrospective and prospective.
③ AVF stenosis was defined as blood flow less than 500 ml/min, which could not satisfy the requirement of dialysis. Technical success was defined as residual stenosis less than 30%.
④ There was no restriction with regard to publication status; however, language was confined to English and Chinese.
⑤ Selected studies contained control and HPB groups.
⑥ Follow-up of patients for at least 6 months.

### Exclusion criteria

① Unrelated research, repeated literature, reviews, case reports, animal experiments, lectures, letters, and anatomical reports.
② Different diagnosis criteria or incomplete data.
③ Studies that investigated other types of vascular access.
④ No other confounding factors

### Literature search strategy

The time of publication was limited to the time the databases were established to November 1, 2020. PubMed, EMBASE, the Cochrane Library, CNKI, WanFang, VIP, SinoMed and other databases were searched to identify literature associated with AVF stenosis and the various treatment approaches. Both subject words and free words were used to retrieve information. The following keywords were used: “autogenous arteriovenous fistula”, “arteriovenous fistula”, “AVF”, “AV fistulae”, “AV fistula”, “arteriovenous fistulas”, “stenosis”, “dysfunction”, “failure and high-pressure balloon”.

### Study selection criteria

A total of 156 articles satisfied the inclusion and exclusion criteria, of which 70 articles were duplicates. Sixty-eight articles were excluded after reading the titles and abstracts; the full text of the remaining 18 articles were evaluated. Ultimately, 9 articles [5–13] comprising 475 patients were included in the current meta-analysis (Figure 1). Characteristics of all included studies are summarized in Table 1. Among the 9 articles, 2 were retrospective cohort studies, 4 were prospective cohort studies, and 3 were randomized controlled trials (RCTs).

**Table 1.** Characteristics of the studies included in the meta-analysis.

| Author, year | Study design | Sample size | Gender (M/F) | Age (year) | Pressure (Atm) | Outcome of interest |
| --- | --- | --- | --- | --- | --- | --- |
| Minli Zhu, 2013 | RCS | HPB: 36<br>CB: 40 | HPB: 22/14<br>CB: 24/16 | HPB: 55. 12 ± 6. 74<br>CB:54.37 ± 6.42 | HPB: 13.14±0.68<br>CB:15.82±0.76 | patency rate (3 month), technical success rate, blood and diameter of AVF, etc. |
| Geng Wang, 2016 | PC | HPB: 20<br>CB: 20 | HPB: 12/8<br>CB: 6/14 | HPB: 60.8±1.95<br>CB: 59.5±2.01 | HPB:15.4±0.84<br>CB:13.2±0.60 | postoperative patency rate (3,6 month), technical success rate, location of AVF, etc. |
| Bisi Wang, 2016 | RCS | HPB: 8<br>CB: 13 | 8/13 | 64.8 | Un-report | postoperative patency rate (3 month), technical success rate, location and residual stenosis of AVF, etc. |
| Guangxing Lu, 2020 | PC | HPB: 39<br>CB: 39 | HPB: 24/15<br>CB: 23/16 | HBP: 52. 36 ± 9. 84<br>CB: 53. 17 ± 11.38 | HPB:16.57±0.58<br>CB:13.29±0.43 | postoperative patency rate (6 month), technical success rate, pressure and time of angiography, etc. |
| Qiongxiang Liang,2020 | PC | HPB: 20<br>CB: 20 | 24/16 | 30~38 | HBP:15.4±0.84<br>CB:13.20±0.60 | postoperative patency rate (3,6 month), technical success rate, pressure of dilation, etc. |
| Qing He, 2017 | PC | HPB: 20<br>CB: 20 | HPB: 11/9<br>CB: 12/8 | HPB: 71<br>CB: 60 | Un-report | postoperative patency rate (3,6,12 month), technical success rate |
| Pasteur Rasuli, 2015 | RCT | HPB: 20<br>CB: 18 | HPB: 11/9<br>CB: 11/7 | HBP: 70<br>CB: 61 | Un-report | postoperative patency rate (3,6,12 month), technical success rate, etc. |
| Syed Arafat Aftab, 2014 | PC | HPB: 35<br>CB: 36 | HBP: 25/10<br>CB: 24/12 | HBP: 57.6 ± 11.58<br>CB: 62.5±10.84 | HBP:21.69±5.38<br>CB:8.17±1.6 | postoperative patency rate (6 month), technical success and complications of angiograph |
| Koki Wakamoto, 2018 | RCT | HPB: 37<br>CB: 32 | HPB: 23/14<br>CB: 20/14 | HPB: 69 ± 10<br>CB: 71 ± 13 | Un-report | postoperative patency rate (3,6,12 month), factors regard to AVF stenosis |

RCS: Retrospective cohort study, PC: Prospective cohort, RCT: Randomized controlled trial, HPB: High-pressure balloon, CB: Conventional balloon, M: male, F: Femal

**Figure 1.**
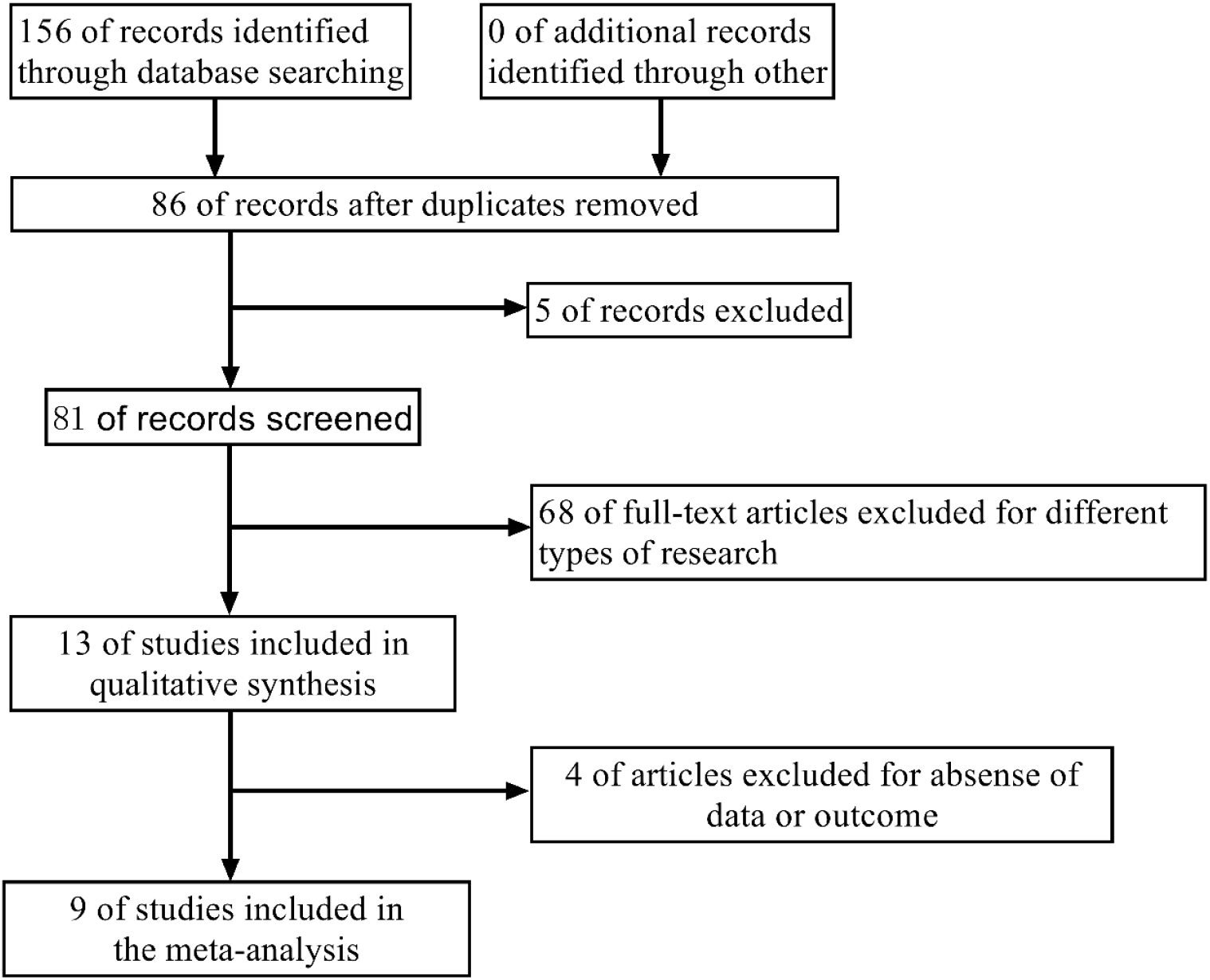
Flowchart of article screening and selection process

### Literature quality assessment and data extraction

Two reviewers independently evaluated the quality of the included studies. Any discrepancies were resolved through a discussion, and a third researcher would make a decision if it was necessary. The quality of each cohort study was assessed using the Newcastle-Ottawa Scale: the selection of study groups, comparability of groups, and outcome of interest. A high-quality study was described as one with a quality score of ≥7. The Cochrane tool was used to evaluate the quality of randomized studies. The score of each study is presented in Table 2 and 3. References from the selected studies were also searched.

**Table 2.** The Cochrane risk of bias tool for assessing the quality of randomized controlled trials included in the meta-analysis.

| RCTs | Random Sequence Generation | Allocation Concealment | Blinding of Participants and Personnel | Blinding of Outcome Assessment | Incomplete Outcome Data | Selective Reporting | Other Biases |
| --- | --- | --- | --- | --- | --- | --- | --- |
| Pasteur Rasuli, 2015 | Unclear | Low risk | Low risk | Unclear | Low risk | Low risk | Low risk |
| Koki Wakamoto, 2018 | Low risk | Low risk | Unclear | High risk | Low risk | Low risk | Low risk |

**Table 3.** The Newcastle-Ottawa Scale for assessing the quality of cohort studies in the meta-analysis*.

| Cohort study | Selection |  |  |  | Comparability of Cohorts | Outcome |  |  | Total Quality Scores |
| --- | --- | --- | --- | --- | --- | --- | --- | --- | --- |
|  | Representativeness of the Exposed Cohort | Selection of the Nonexposed Cohort | Ascertainment of Exposure | Outcome of Interest Was Not Present at Start of Study |  | Assessment of Outcome | Follow-up Period Long Enough for Outcomes to Occur | Adequacy of Follow-up Evaluation of Cohorts |  |
| Minli Zhu 2013 | 1 | 1 | 1 | 1 | 1 | 1 | 1 | 1 | 8 |
| Geng Wang 2016 | 1 | 1 | 1 | 1 | 2 | 1 | 1 | 1 | 9 |
| Bisi Wang 2016 | 1 | 1 | 1 | 1 | 1 | 1 | 1 | 1 | 8 |
| Guangxing Lu, 2020 | 1 | 1 | 1 | 1 | 2 | 1 | 1 | 1 | 9 |
| Qiongxian Liang, 2020 | 1 | 1 | 1 | 1 | 1 | 1 | 1 | 0 | 8 |
| Qing He, 2017 | 1 | 1 | 1 | 1 | 1 | 1 | 1 | 1 | 8 |
| Syed Arafat Aftab, 2014 | 1 | 1 | 1 | 1 | 2 | 1 | 1 | 1 | 9 |

\*A study can be awarded a maximum of 1 point for each numbered item within the “Selection” and “Outcome” categories. A maximum of 2 points can be awarded for comparability.

Data extracted from the studies included: author, title, publication date, literature source and other general information; research object, intervention measures, study design, follow-up time, outcome indicators, etc.

### Assessment of heterogeneity and publication bias

Heterogeneity was assessed using the I^2^ statistic and P value; I^2^ statistic values of <25%, 25% to 50%, and >50% were considered as low, moderate, and high heterogeneity, respectively. An I^2^ value >50% (p<0.05) denoted significant heterogeneity across the included studies. A potential publication bias was estimated using a funnel plot (Figure 3). The funnel plot was roughly symmetrical, suggesting that the publication bias was not evident.

### Statistical and sensitivity analyses

Meta-analyses were performed on the outcomes based on fixed and random effects models to estimate the odds ratios (ORs) and 95% confidence intervals (CI) due to heterogeneity of studies. Furthermore, subgroup analyses based on follow-up time were conducted. Sensitivity analyses were also performed to test the reliability of the results by removing one study at a time and repeating the meta-analysis. Based on the sensitivity analyses, a study by Syed Arafat Aftab had a considerable influence on the results at 6 months after operation. The article was removed and a meta-analysis (random effects model) was conducted again, and the results revealed an OR value of 0.33, 95% CI 0.15-0.75, P = 0.008. Analyses were performed with RevMan 5.3.

## Results

Nine studies comprising 475 patients with AVF stenosis were included in the meta-analysis. The characteristics of the patients are presented in Table 1. Among the 9 articles, 2 were retrospective cohort studies, 4 were prospective cohort studies, and 3 were RCTs. AVF stenosis was reported in all studies. The pooled results revealed that stenosis rate of AVFs treated with HPB was significantly lower than that of AVFs treated with CB at 3 months (OR= 0.37, 95% CI 0.21 to 0.67, p<0.001) and 6 months after operation (OR= 0.33, 95% CI 0.15 to 0.75, p=0.008). In addition, the technical success rate of the HPB groups was high (OR= 0.14, 95% CI 0.05 to 0.35, p<0.001). However, no significant difference was observed between the experimental and control groups at 12 months after operation (OR= 0.61, 95% CI 0.29 to 1.25, p=0.18). The results are presented in Figure 2. A funnel plot representing publication bias of the studies is presented in Figure 3; the funnel plot was symmetrical, which indicated that there was a slight publication bias.

**Figure 2.**
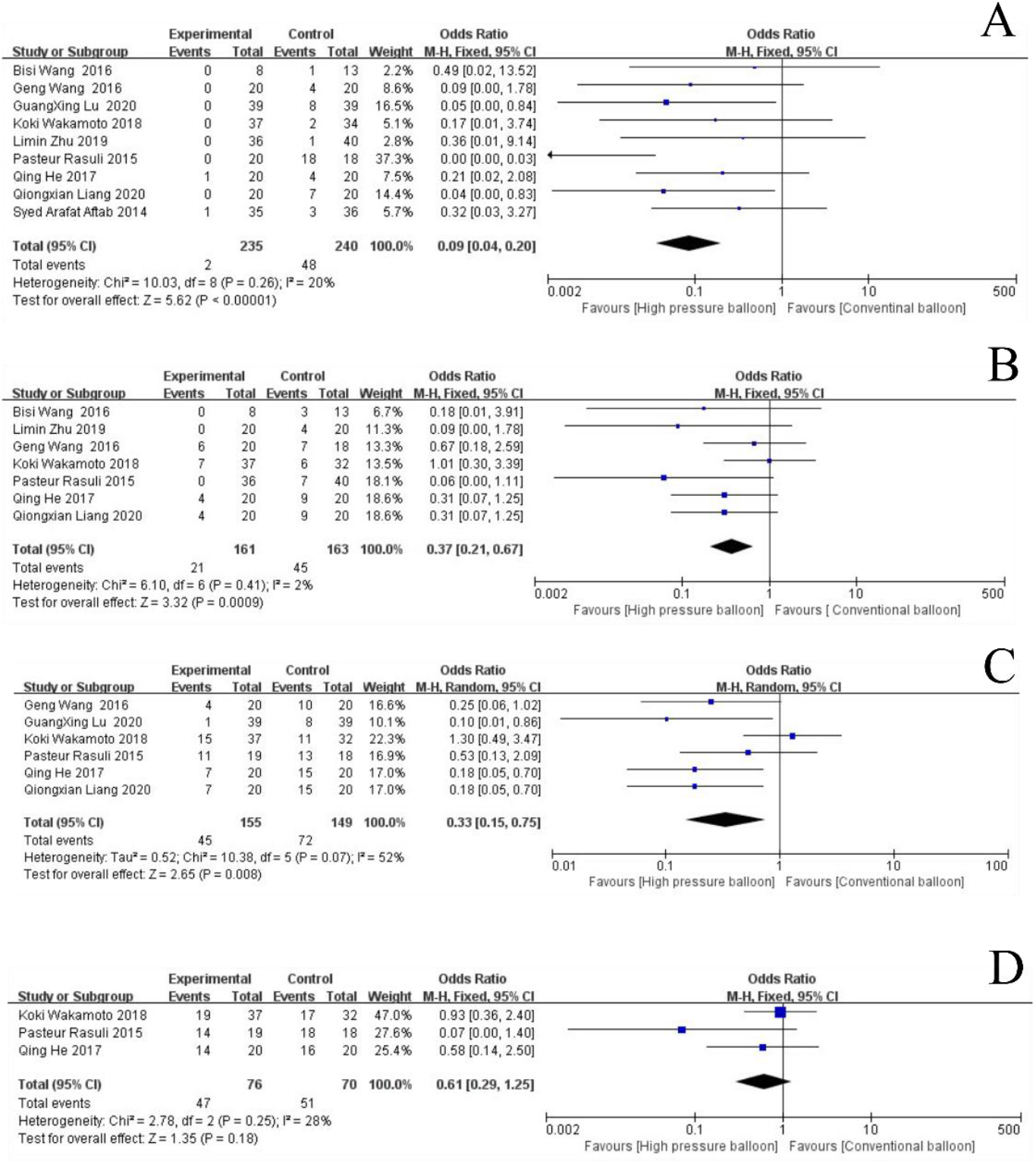
Forest plot for (A) technical success rate and patency rate at 3 month (B), 6 month (C) and 12 month (D)

**Figure 3.**
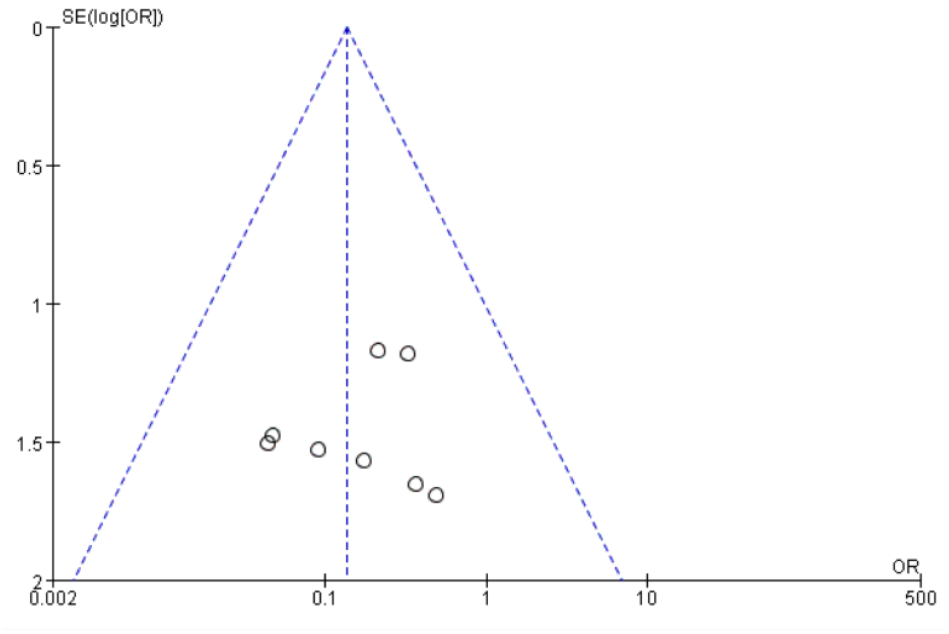
Funnel plot is roughly symmetrical, indicating that publication bias is not obvious

## Discussion

With the prolongation of the survival time of patients with dialysis, salvaging stenotic AVF to extend their patency time is equally important as creating them. According to the consensus of vascular access experts in China in 2019 [14], indications of surgical intervention for AVF stenosis predominantly include the following aspects: blood flow < 500 ml/min (does not satisfy blood flow requirements for dialysis), high static pulse pressure; puncture complications lead to decrease in dialysis adequacy. However, the type of balloon that should be used to treat AVF stenosis remains undetermined. The conclusions of previous studies are also inconsistent, which presents a challenge that requires urgent resolution. In addition, the latest Kidney Disease Outcomes Quality Initiative guidelines [15] recommend HPB as a first choice. Therefore, a comprehensive search of the medical databases and statistical analyses were conducted. We attempted to identify the type of balloon that could be suitable for the treatment of AVF stenosis to provide a theoretical basis for treatment. The present study is the first to conduct a meta-analysis on the potential application of HPB for the treatment of AVF stenosis.

Previous studies have revealed that HPB is superior to BA in the treatment of coronary atherosclerotic stenosis [16]. Furthermore, a few studies have demonstrated that dilation of AVF stenosis requires higher pressure than that of atherosclerotic stenosis [17], which is consistent with the results of the present meta-analysis (at 3 months and 6 months after operation). However, the results of the present meta-analysis suggest that the long-term effect (12 months after operation) on the HPB group is not satisfactory.

The burst pressure of HPB could up to 35–45 atm [18]. The maximum pressure of HPB applied at the center where the author is located was nearly 20 atm. HPB not only exhibits a high burst pressure but also exhibits no change in compliance before attaining the maximum pressure, which confers the advantage of curing lesions resistant to CB [19]. In addition, one of the factors associated with AVF stenosis is arterial calcification. A few studies suggested that HPB was an effective treatment for calcified arteries [20]. Therefore, HPB can exhibit a higher technical success rate, which is consistent with the results of the present meta-analysis. Nevertheless, HPB also has negative effects due to its high pressure, which may cause vascular endothelial damage and it is more likely to cause vascular restenosis in subsequent dialysis [21]. Schiele et al. demonstrated that moderate balloon inflation pressure could benefit patients with restenosis [22]. This mechanism could explain why the long-term effect of HPB is not satisfactory.

The present study had a few limitations with regard to the quality of the published studies included in the meta-analysis. Although most of the studies included in this analysis were prospective and randomized, the quality of certain studies was unsatisfactory (largely due to the imperfect blinding methods).

In conclusion, HPB is a preferred choice of AVF stenosis, which is consistent with the most recent guidelines. However, further studies should be conducted to investigate the mechanisms of restenosis after angiography. The efficacy of other balloons for the treatment of AVF stenosis requires further studies.

## Data Availability

This data that support the findings of this study are openly avaliable in corresponding author upon reasonable request.

## Author Contributions

Yu Li and Wenhao Cui. searched the literature, analyzed the extracted data, and wrote manuscript. Jukun Wang and Chao Zhang. participated in discussion. Tao Luo. supported writing manuscript. All authors agreed to publish the manuscript. All authors have read and agreed to the published version of the manuscript.

